# Development of a Children and Young People’s Informed Framework for the Design, Delivery and Evaluation of a Patient and Public Involvement, Engagement and Participation Group: A Service Development Project

**DOI:** 10.64898/2026.10.21.26366171

**Authors:** Jane Chudleigh, Gemma Jones, Lucy Pickard, Manon Deverese, Pru Holder, Anisa Singh, Nandeeta Samad, Abigail Rose, Jennifer Preston, Ingrid Wolfe

**Author notes:** **Author contributions:** Conceptualization, JC, GJ, MD, IW, LP; methodology, JC, GJ, MD, LP; data collection, JC, MD, PH, NS; formal analysis, JC, PH; writing—original draft preparation, JC; writing—review and editing, JC, GJ, MD, PH, IW, AS, NS, AR, JP, LP; project administration, JC, GJ, MD, AS, NS, LP; funding acquisition, JC, GJ, MD, IW, AS, NS, LP. All authors have read and agreed to the published version of the manuscript.

## Abstract

**Background:** Patient and Public Involvement, Engagement and Participation (PPIEP) is increasingly recognised as an essential component of high-quality health research, ensuring research is relevant, acceptable and meaningful to the communities it aims to serve. PPIEPP activities remain predominantly on adults; focused children and young people (CYP) are often underrepresented during the design and implementation of research studies that affect them. It is therefore vital their direct experiences, priorities and preferences are captured.

**Objective:** To develop a CYP-informed approach to meaningful involvement to guide the future development of a CYP PPIEP group.

**Design:** A service development project using qualitative methods (online focus groups incorporating Mentimeter) to explore perspectives and experiences of involvement, engagement and participation in research with CYP with long-term health conditions and to co-develop key elements of a future CYP PPIEP group.

**Results:** Thirteen CYP (7 males and 6 females) aged 12-16 years (median 14 years) from a range of ethnic backgrounds and long-term health conditions were involved in n=4 online focus groups. CYP expressed a preference for flexible meeting options outside school hours, with hybrid delivery options to improve accessibility, alongside advance notice, reminders and clear meeting agendas. CYP also expressed the importance of researchers creating an environment where CYP feel listened to, respected and able to contribute openly. CYP provided recommendations on recruitment strategies for CYP PPIEP groups, information needs prior to joining, motivations for participation, the inclusion of diverse CYP groups, creating a safe environment for sharing views, and training requirements to support meaningful involvement.

**Conclusion:** CYP identified key principles to inform the development of a CYP PPIEP group. These principles were mapped against the UK Standards for Public Involvement and National Institute for Health and Care Research (NIHR) guidance to develop a CYP-informed framework for the design, delivery and evaluation of a CYP PPIEP group.

## 1.0 Introduction

Patient and Public Involvement, Engagement and Participation (PPIEP) is increasingly recognised as an essential component of high-quality health research, ensuring that research is relevant, acceptable and meaningful to the communities it aims to serve(1). The UK Policy Framework for Health and Social Care Research sets out the key principles expected to underpin the management and conduct of health and social care research across the UK, with Principle 4 specifically highlighting the importance of involving patients, service users and the public throughout the research process. In particular, the Framework emphasises that public involvement should extend across areas such as research design, management, conduct and dissemination, rather than being limited to a single stage of the research process(2). However, PPIEP activities are often focused on adults, meaning the views and experiences of children and young people (CYP) are underrepresented(3–5). Article 12 of the United Nations Convention on the Rights of the Child clearly states that children should have the opportunity to be heard in decisions that directly affect them(6). The Lundy Model of Child Participation describes four criteria necessary to ensure Article 12 is implemented meaningfully, transparently and with integrity. These are, ‘Space’, providing CYP with a safe environment to express their views; ‘Voice’, supporting CYP to communicate their views; ‘Audience’, ensuring CYP views are listened to by those responsible for decisions; and ‘Influence’, ensuring CYP views are taken seriously and, where appropriate, inform decision(7). Despite this, CYP may be structurally and/or practically denied agency, or perceived as lacking decision-making competence in research(8). Consequently, assent may be reduced to a tokenistic process rather than recognised as a genuine reflection of children’s evolving autonomy and right to accept or decline participation(9, 10). There is also no universal agreement on the age at which CYP should be considered able to participate independently in PPIEP activities(11). In addition, children of the same age may demonstrate varying levels of maturity and understanding and therefore age, context and competence all contribute to their decision-making(11).

Currently, there are two commonly used models of CYP PPIEP; the first, often referred to as young people’s advisory groups (YPAGs), consist of ongoing, sustainable groups that can involve CYP across multiple research projects. The second consist of CYP PPIEP groups set up for individual research projects, with activities often concentrated within the boundaries of the funded research lifecycle rather than sustained across projects(12). The project-specific model may be particularly problematic for CYP, as continuity of relationships and opportunities to develop expertise may be disrupted by the time-limited nature of research funding(13).

While consideration has been given to involving CYP in decision-making in research(10, 14), there is less robust evidence regarding how CYP can be meaningfully involved as PPIEP members to enable them to shape research from the outset(13, 15). A scoping review found ambiguities in the quality of reporting of PPIEP with CYP and variation regarding their level of involvement(13). A review of how CYP involvement was reported in the design and conduct of child health research also found variability in the way CYP PPIEP was reported, with few studies formally evaluating PPIEP involvement(16). Similarly, a rapid umbrella review found that while there is consistency in the methods used (such as arts-based methods like drawing and photovoice especially with children less than 12 years of age) and stages at which CYP are involved in PPIEP activities, there is a lack of consistent reporting and evaluation of the impact of PPIEP with CYP(15).

The UK Standards for Public Involvement(17) consist of six standards to provide clear, concise statements of effective public involvement against which improvement can be assessed. While it is stated these are adaptable to different situations, none of the 40 pilot projects used to refine and better understand the experience of implementing the standards focussed on CYP. The National Institute for Health and Care Research (NIHR) guidance provides practical recommendations to support meaningful CYP involvement in research, including involving CYP from the earliest stages of research, providing appropriate support and training, enabling inclusive participation and demonstrating the impact of their contribution(18). However, it does not consider how this involvement is evaluated.

Existing literature has largely focused on how researchers can involve CYP, rather than assessing and supporting the quality of PPIEP specifically from a CYP perspective. The aim of this service development project was to work with CYP to inform the development of a future CYP PPIEP group. The resulting themes and principles were subsequently mapped against the UK Standards for Public Involvement(17) and NICE guidance(18) to inform the development of a CYP-informed PPIEP framework intended to support the development, delivery and evaluation of a meaningful CYP PPIEP group.

## 2.0 Materials & Methods

### 2.1 Aim

To develop a CYP-informed approach to meaningful PPIEP to guide the future development of a CYP PPIEP group.

### 2.2 Participants

CYP aged 10-17 years with a long-term health condition were invited to participate. Recruitment occurred via two routes: previous involvement in research and/or PPIEP activities, and through an outpatient clinic at a tertiary care hospital. CYP and their parents were provided with an information sheet outlining the project and were given at least 48 hours to consider whether they wished to participate. Written or electronic consent was obtained from a parent, alongside written or electronic assent from the CYP prior to the focus groups.

### 2.3 Data Collection

Four focus groups were convened according to participant availability; each participant attended two focus groups. Each focus group was supported by two facilitators (JC and MD, PH or NS). Emphasis was placed on creating a safe and inclusive environment where CYP could share their experiences, priorities, suggestions and views. To support this, Mentimeter, an interactive audience response platform, was used during the focus groups as a data collection tool to facilitate engagement and provide all participants with an opportunity to contribute their perspectives. The use of Mentimeter was also considered appropriate for the age range of participants. It also provided an accessible way for them to share their views, including where they may have felt less comfortable contributing verbally within the group. At the beginning of each focus group, participants were introduced to Mentimeter through a small number of test questions. This provided an opportunity for CYP to familiarise themselves with the functionality, explore how responses were submitted, and build confidence in using the tool before moving on to the focus group activities.

A range of question formats were used to capture different types of responses, including multiple-choice questions, open-ended responses, word clouds, ranking activities, and rating scales. This enabled CYP to share individual perspectives, prioritise key themes, and reflect on areas where consensus or differing views emerged. The combination of structured and open-ended activities provided opportunities for both quantitative-style responses (e.g., rankings and scales) and richer qualitative insights through written comments and discussion. Responses generated through Mentimeter from the first round of focus groups were used to inform the subsequent discussion and explore participants’ views in greater depth during the second round of focus groups.

The transcription function in Microsoft Teams was used to produce verbatim transcripts of the focus groups that were checked for accuracy by the focus group facilitators. Comments in the chat were also downloaded and included in the analysis.

### 2.4 Data analysis

Responses collected via Mentimeter were downloaded and analysed using a mixed descriptive approach. Closed-ended responses were summarised using simple descriptive statistics, primarily frequencies. Open-ended responses were coded inductively, with related codes grouped into overarching themes (JC) that were checked by a second member of the research team (PH); no disagreements arose. Frequencies were reported where appropriate to indicate the number of participants identifying particular issues. The resulting themes were collaboratively mapped (JC, PH) to the UK Standards for Public Involvement in Research(17) and NIHR guidance(18) to inform the development of a CYP-informed PPIEP framework.

## 3.0 Results

Thirteen CYP (n=7, 54% male, n=6, 46% female, aged 12-16 years, median 14 years) were recruited. Four focus groups were held during 01 June 2026-30 June 2026. The first round of focus groups (focus groups 1 and 2) involved 12 of the 13 CYP and explored how the proposed CYP PPIEP group should operate, including participants’ views on its structure, purpose, and ways of working. The second round of focus groups (focus groups 3 and 4) also involved 12 of the 13 CYP and explored recruitment, accessibility, inclusion, psychological safety, and training needs. One participant who attended the first round was unable to attend the second round, while a different participant attended the second round but not the first due to availability. Therefore, the denominator used to calculate percentages reported within themes is 12 CYP.

CYP identified as being from a range of ethnic backgrounds, including Black, Black British, Caribbean or African (n=7, 54%), White (n=3, 23%), and Asian or Asian British (n=3, 23%). CYP also reported experience with a range of paediatric specialties, including haematology (n=5, 39%), endocrinology (n=2, 15%), gastroenterology (n=2, 15%), neurology (n=1, 8%), oncology/neurology (n=1, 8%), and participants who preferred not to disclose this information (n=2, 15%).

### 3.1 Theme 1: Meaningful Partnership

CYP were asked what would make participation in a PPIEP group feel meaningful. Many responses (n=8, 67%) emphasised the importance of seeing tangible impact and progress from their involvement “If [PPIEP] could lead to better results/change in the way which things are done”. CYP wanted to know how their contributions could influence a project and lead to meaningful improvements or changes.

CYP wanted to be listened to “Children have voices and should be and can be listened to.” CYP (n=4, 33%) expressed that if their views were not listened to or were altered in ways that no longer reflected their original meaning, this might negatively affect their experience of PPIEP involvement. These CYP expressed concerns about contributing ideas that were subsequently ignored or modified “…answers being changed slightly through an adult lens which defeats the point”.

Participants identified the opportunity to make a difference and contribute to improving their own and the lives of others (n=4, 33%) and having their voices heard and valued (n=3, 25%) as important motivating factors “Being heard and being around people that are going through the sort of same thing.”

### 3.2 Theme 2: Inclusive recruitment and accessibility

Participants identified a range of approaches for advertising a future CYP PPIEP group. Digital platforms and social media were the most frequently suggested recruitment routes (n=8, 67%), with participants highlighting platforms including Instagram, TikTok, Facebook, YouTube, and social media groups as ways to reach CYP. Schools and education settings were also identified as possible routes for reaching CYP (n=2, 17%), alongside healthcare settings such as hospitals (n=1, 8%). Community-based approaches, including local youth centres and libraries, were also suggested (n=2, 17%), as were printed materials such as leaflets (n=1, 8%).

Participants expressed a preference for flexible approaches to how meetings are delivered; time-related factors were considered the main barrier to attending PPIEP meetings (n=11, 92%). A hybrid format, combining online and in-person participation, was suggested (n=6, 50%). Four (33%) participants preferred meetings to be held online, while one (8%) participant preferred in-person meetings. A hybrid approach was also considered to accommodate CYP with health conditions, school commitments, or other barriers that may prevent them attending in person, “Some people might prefer being anonymous and some people might be more comfortable with like being there with other people.”

Most participants preferred a regular meeting schedule, with monthly meetings identified as the most popular option (n=9, 75%). Three (25%) participants preferred meetings every two months, while one (8%) participant preferred meetings every three months. Additional suggestions included allowing flexibility depending on the needs of the project and ensuring meetings were arranged at appropriate intervals to maintain engagement without becoming burdensome.

Most participants felt that meetings should last approximately one hour (n=9, 75%), suggesting that shorter, focused sessions may be more appropriate to maintain engagement and avoid meetings feeling overwhelming. Three (25%) CYP provided alternative suggestions, including adapting meeting length depending on the activity or topic being discussed.

Participants identified several ways researchers could make it easier for CYP to attend PPIEP meetings. The most frequently identified theme was the importance of flexibility in meeting arrangements (n=8, 67%). CYP highlighted the need for flexible and convenient meeting times, consideration of school commitments, scheduling meetings around school holidays where possible, ensuring participants were available, and avoiding last-minute changes.

Eight (67%) of CYP expressed that CYP who were neurodivergent, autistic, or who experienced attention difficulties, or other communication needs may require additional support to participate effectively. Five (42%) CYP thought that younger children may find it more difficult to be part of a PPIEP group. Social anxiety, shyness, and confidence were also identified as potential barriers to participation (n=4, 33%). Participants considered that adaptations and additional support may enable full participation and suggested allowing more time to process information, breaking activities into smaller steps, providing clear explanations, and considering different communication needs “…they might not understand…they will need more time to process information…break it down into steps.”

### 3.3 Theme 3: Capacity building and personal development

Participants identified a range of opportunities that would make involvement in a PPIEP group valuable for their personal, educational and professional development. Understanding more about research was identified as the highest priority in terms of learning opportunities, with n=7 (58%) of CYP ranking this as their highest priority. Other learning priorities included understanding how the NHS works (n=2, 17%) and understanding how research is funded (n=2, 17%).

Experience of undertaking research (e.g., opportunity to be a peer researcher) was identified as an potential future benefit by CYP (n=5, 42%). Participants also highlighted the possible benefits of PPIEP involvement for future education “…I do sociology too so it could be helpful” and employment “…it can give me experience if I am going to work in the research sector” (n=4, 33%).

Opportunities to gain experience and develop skills were highlighted (n=3, 25%) by CYP. Gaining confidence (n=3, 25%), and experience, knowledge (n=4, 33%) were also identified as important motivators for continued involvement “…a way to get experience of "adult life" without being thrown in the deep end” and communication skills.

### 3.4 Theme 4: Recognition and reciprocal benefit

CYP were keen to understand the purpose of the group “what your input would go towards, type of research, the aim e.g., helping people” (n=5, 42%) and what they would gain from involvement, including opportunities to develop skills and experiences (n=5, 42%) and payment (n=3, 25%) “…payment while learning practically” before deciding whether to be involved in a CYP PPIEP group.

CYP highlighted the importance of feeling that their time and contributions were valued and appreciated. Payment, rewards, recognition and opportunities were also the most frequently identified factors (n=7, 58%) to ensure sustained involvement in a CYP PPIEP group.

CYP also highlighted the potential reciprocal benefits to themselves “difference to our lives…add to CV….making a difference to others” and researchers “…young people have more helpful things to say than expected.”

### 3.5 Theme 5: Safe, engaging and supportive environment

The experience of being involved was also viewed as important. CYP highlighted that PPIEP activities should be enjoyable, engaging, and provide opportunities to work collaboratively with others (n=7, 58%). Examples included creating something together, such as a video, trying new activities, and having a positive group experience where participants felt comfortable contributing.

CYP highlighted that confidence in sharing opinions “…comes with time” and depends on feeling comfortable within the group. Building trust and familiarity was identified as important in helping CYP feel able to contribute openly (n=4, 33%). One CYP highlighted the importance of being part of a smaller group where CYP may feel more comfortable contributing (n=1, 8%) “…eight ish people max, or less than 10.”

CYP also highlighted the importance of having choice and not feeling pressured to share before they were ready (n=2, 17%). Participants suggested that opportunities to contribute should be encouraged but remain optional, allowing CYP to participate at their own pace. Additional factors that may support comfort in sharing included anonymity where appropriate (n=1, 8%) and opportunities for face-to-face interaction to help build relationships within the group (n=1, 8%).

While n=6 (50%) of CYP indicated that the existing format worked well, they also shared ideas for making sessions fun, accessible and enjoyable. Interactive activities were the most frequently identified approach (n=8, 67%), with CYP highlighting the value of games, interactive tasks, and tools such as Mentimeter, which they felt encouraged participation and made sessions more engaging “Menti is really good”. Creating opportunities for everyone to contribute and “…express themselves easily” was also considered important (n=3, 25%). CYP also valued sessions that felt conversational and relaxed (n=4, 33%).

The importance of having private and confidential routes for providing feedback was also mentioned (n=5, 42%). CYP suggested approaches such as email, private messages, and direct communication with facilitators (adult researchers) as ways to share concerns safely. Anonymous feedback options were also highlighted (n=2, 17%), including anonymous messages and the ability to provide feedback without being identified. Participants also suggested using familiar messaging platforms, such as WhatsApp, to support accessible communication (n=2, 17%), although this would need to follow relevant organisational safeguarding, consent and data-protection requirements for CYP. Other approaches included opportunities for one-to-one conversations with facilitators (n=1, 8%) and involving parents, especially for younger children (n=1, 8%).

Participants also valued positive relationships and opportunities to connect with others, including meeting researchers (n=2, 17%).

### 3.6 Theme 6: Transparency, feedback and impact

Lack of visible progress, outcomes, or feedback were identified as potential barriers to the success of a CYP PPIEP group (n=6, 50%). CYP highlighted the importance of understanding how their input contributed to improving outcomes. CYP felt that involvement could be perceived as a poor use of their time if they did “Not see a change…” resulting from their contributions, were “…not kept updated throughout…”, or perceived that the project had not achieved a meaningful outcome “…if at the end of the day, the project is not acceptable or not good enough to be published”.

Participants emphasised the importance of seeing how their contributions influenced research. Most participants (n=9, 75%) stated they would prefer to receive updates about the impact of their involvement monthly. When asked how they would like to receive updates, participants identified a range of preferred approaches. The most frequently selected option was receiving updates at the start of each meeting (n=7, 58%), followed by short emails or newsletters (n=6, 50%). Participants also valued opportunities to attend presentations where findings could be shared (n=5, 42%), as well as direct communication from researchers about changes made as a result of their input (n=4, 33%). CYP also highlighted the importance of being able to communicate directly with the group or facilitators (n=4, 33%), including through messages or other direct contact methods. Participants highlighted the importance of regular communication between meetings, including reminders to support ongoing engagement “Whenever there’s anything to update about if that’s an update two days in a row then nothing for two months or once a month.”

The six interconnected principles derived from the focus group findings were mapped against the UK Standards for Public Involvement in Research(17) and the more recent NIHR guidance for involving CYP as advisors in research(18) (Table 1) to develop a CYP-informed framework for the design, development and evaluation of a CYP PPIEP group (Figure 1).

**Figure 1:**
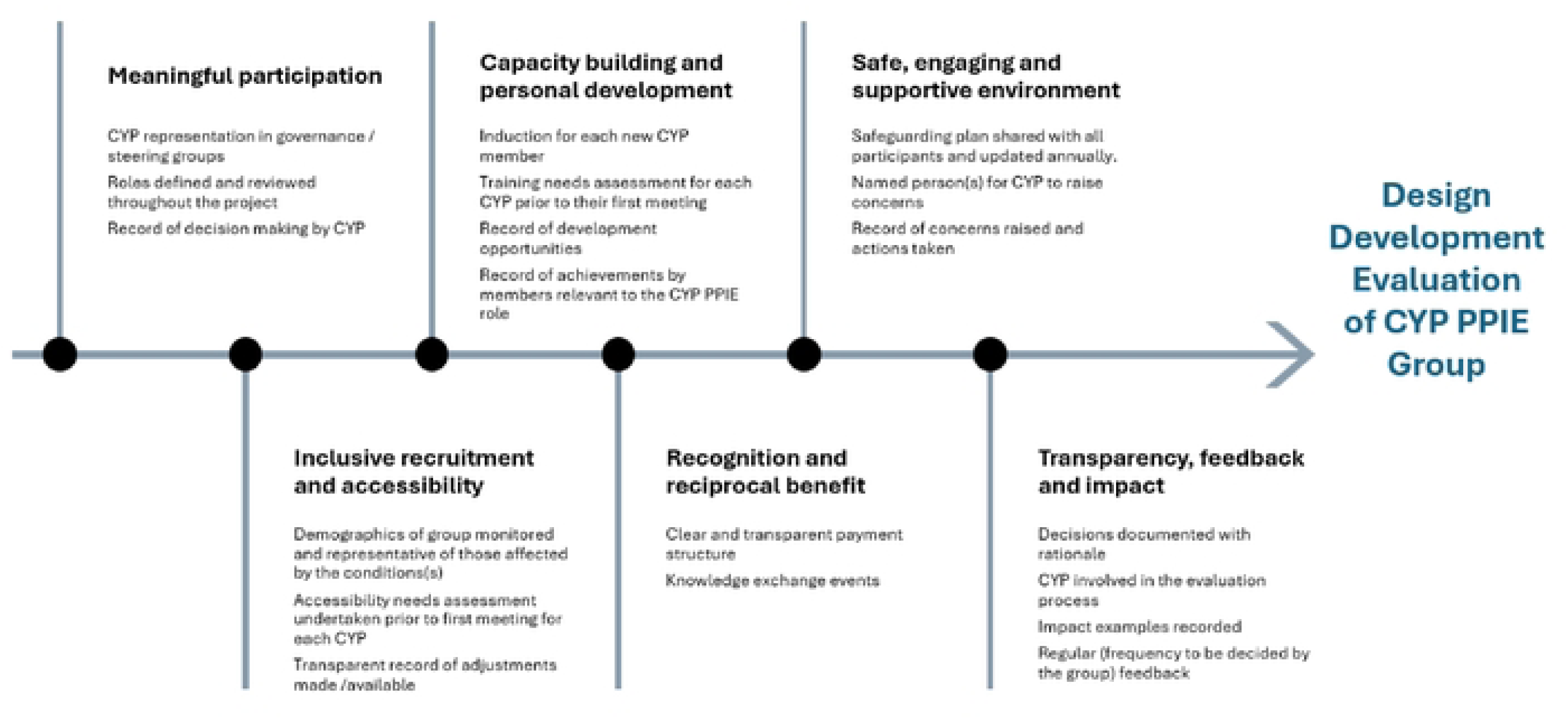
Proposed CYP PPIEP Framework

**Table 1:**
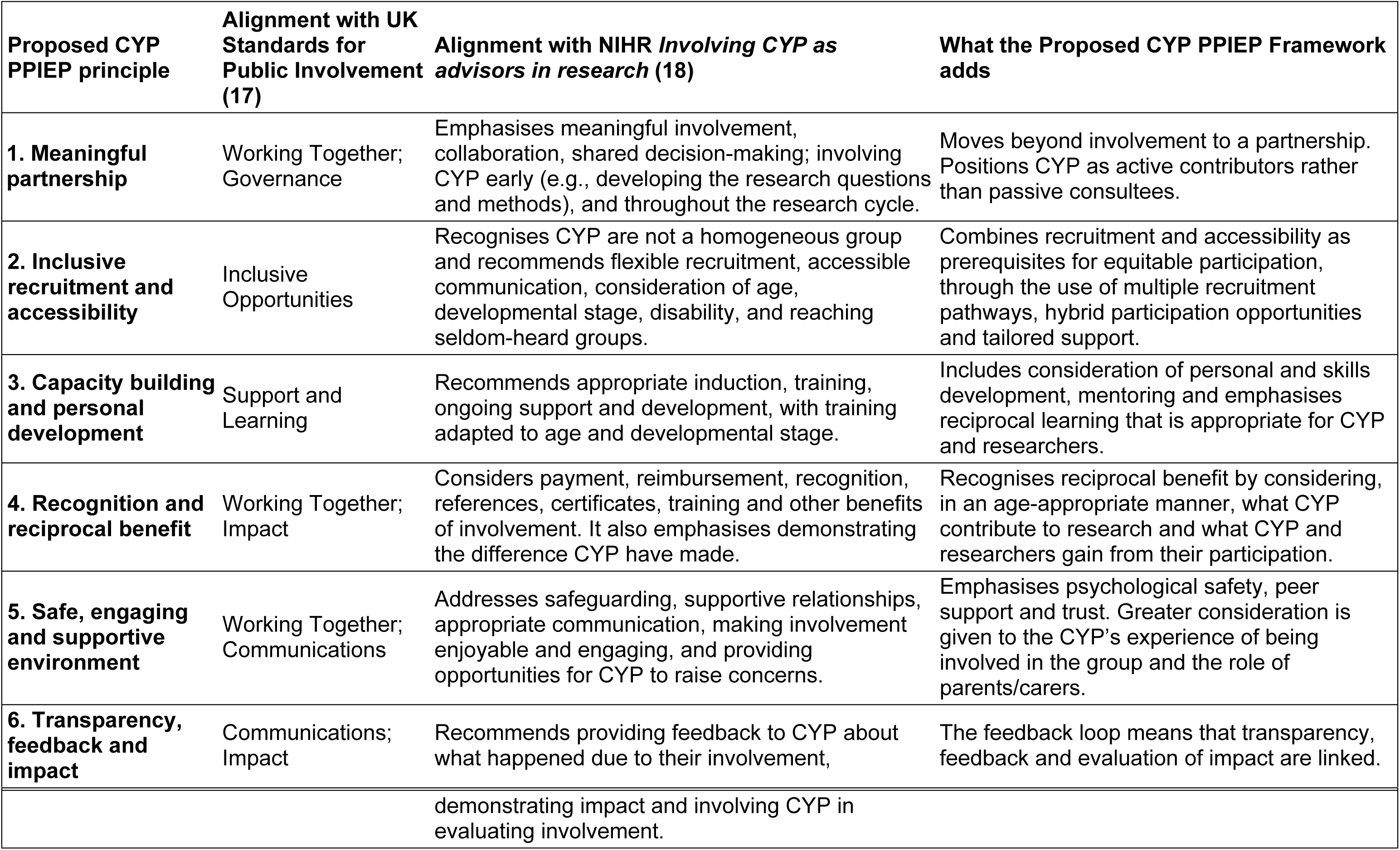
CYP PPIEP principles mapped against the UK Standards for Public Involvement in Research (17) and the more recent NIHR guidance for involving CYP as advisors in research (18)

| **Proposed CYP PPIEP principle** | **Alignment with UK Standards for Public Involvement (17)** | **Alignment with NIHR *Involving CYP as advisors in research* (18)** | **What the Proposed CYP PPIEP Framework adds** |
|---|---|---|---|
| **1. Meaningful partnership** | Working Together; Governance | Emphasises meaningful involvement, collaboration, shared decision-making; involving CYP early (e.g., developing the research questions and methods), and throughout the research cycle. | Moves beyond involvement to a partnership. Positions CYP as active contributors rather than passive consultees. |
| **2. Inclusive recruitment and accessibility** | Inclusive Opportunities | Recognises CYP are not a homogeneous group and recommends flexible recruitment, accessible communication, consideration of age, developmental stage, disability, and reaching seldom-heard groups. | Combines recruitment and accessibility as prerequisites for equitable participation, through the use of multiple recruitment pathways, hybrid participation opportunities and tailored support. |
| **3. Capacity building and personal development** | Support and Learning | Recommends appropriate induction, training, ongoing support and development, with training adapted to age and developmental stage. | Includes consideration of personal and skills development, mentoring and emphasises reciprocal learning that is appropriate for CYP and researchers. |
| **4. Recognition and reciprocal benefit** | Working Together; Impact | Considers payment, reimbursement, recognition, references, certificates, training and other benefits of involvement. It also emphasises demonstrating the difference CYP have made. | Recognises reciprocal benefit by considering, in an age-appropriate manner, what CYP contribute to research and what CYP and researchers gain from their participation. |
| **5. Safe, engaging and supportive environment** | Working Together; Communications | Addresses safeguarding, supportive relationships, appropriate communication, making involvement enjoyable and engaging, and providing opportunities for CYP to raise concerns. | Emphasises psychological safety, peer support and trust. Greater consideration is given to the CYP’s experience of being involved in the group and the role of parents/carers. |
| **6. Transparency, feedback and impact** | Communications; Impact | Recommends providing feedback to CYP about what happened due to their involvement, demonstrating impact and involving CYP in evaluating involvement. | The feedback loop means that transparency, feedback and evaluation of impact are linked. |

The framework therefore provides a practical approach to supporting meaningful, equitable and reciprocal involvement of CYP.

## 4.0 Discussion

This work set out to develop a CYP-informed approach to meaningful PPIEP to guide the future development of a CYP PPIEP group.

CYP in the current project valued knowing that their views could influence research, contribute to positive change, and help improve outcomes for other young people. However, they were aware that their views could sometimes be misrepresented, potentially reducing the impact of their contributions and limiting their influence on decision-making. This idea of children lacking agency(8) or relying on adults’ views as proxies for children’s perspectives has been commonly cited in the literature(19). The findings of this work are consistent with the Lundy Model of Child Participation(7), particularly its emphasis on creating space for children’s voices to be heard. The focus groups provided a supportive space for CYP to express their views and generated valuable perspectives on what meaningful and effective PPIEP could look like in practice. Article 12 of the United Nations Convention on the Rights of the Child places an obligation on signatories not only to listen to CYP but also to act on their views, which aligns with the Lundy Model’s(7) emphasis on audience and influence. In this work, CYP perspectives were used to inform the development of the CYP-informed PPIEP framework, providing a mechanism through which their contributions could influence the research process.

While voices of CYP are often underrepresented in research(3–5), CYP from marginalised backgrounds may face additional barriers to meaningful participation and are consequently further underrepresented(20). Recently a framework has been proposed for PPIEP activities with CYP with rare kidney diseases that includes an evaluative component. However, this framework has a strong focus on recruitment strategies(21), whereas the proposed framework takes a broader approach, considering CYP involvement across the development, delivery and evaluation of research. This extends beyond recruitment to provide a more comprehensive approach to supporting meaningful CYP involvement throughout the research process. CYP involved in the current project represented a range of long-term health conditions, bringing diverse experiences and perspectives. Drawing on these experiences, they identified approaches to support more equitable participation, including multiple recruitment pathways, flexible opportunities for participation and support tailored to CYP’s needs.

The importance of creating a safe space for CYP to share their thoughts and ideas has been emphasised previously(20). In the current project, providing multiple ways for CYP to raise concerns was considered important to ensure that they felt safe, could express honest feedback and that concerns could be acknowledged and addressed appropriately. Previous work has identified ambiguities in the quality of reporting of PPIEP with CYP(13, 15) and a lack of consistent evaluation of the impact of PPIEP with CYP(15). CYP in the current project emphasised the importance of regular feedback loops in maintaining trust and demonstrating that CYP involvement has a meaningful impact.

Although the original intention was to use these perspectives to inform the development of a specific PPIEP group, mapping the resulting data against the UK Standards for Public Involvement(17) and the NIHR guidance for involving CYP as research advisors(18) provided an additional opportunity to synthesise these insights into a broader CYP-informed framework.

The six principles developed through the service development project demonstrated alignment with UK Standards for Public Involvement(17) and the NIHR guidance for involving CYP as research advisors(18). The resulting CYP-informed framework brings these together to provide a structured approach through which the quality of CYP PPIEP can be considered, while retaining the practical considerations identified within existing guidance. The proposed framework provides a structured way of considering the design, development and evaluation of CYP PPIEP groups.

As with any service development project, this work had both strengths and limitations. The work was undertaken with adolescents and therefore the views of younger children have not been represented in the framework. Some CYP had prior research/PPIEP involvement experience, while others were recruited through a tertiary clinic; therefore, their perspectives may differ from those of CYP with no previous involvement or less contact with specialist services. While the number of included CYP was relatively small, they had lived experience of a range of health conditions and were able to bring a breadth of perspectives that could strengthen the relevance and inclusivity of the project. The inclusion of a relatively high proportion of CYP from ethnically marginalised groups also contributed to a diverse range of perspectives and aligned with the project’s commitment to equity, diversity and inclusion. As this work was undertaken as a service development project rather than research, the framework should not be interpreted as a research-derived or empirically validated model, nor as being universally generalisable. Rather, it represents a useful starting point for other teams seeking to establish, develop or evaluate CYP PPIEP groups.

## 5.0 Conclusions

This project demonstrates how a service development activity, when grounded in CYP involvement and considered alongside established standards for public involvement(17, 18), can generate learning with potential wider utility beyond the immediate service context.

Future work could explore the applicability and usefulness of the framework across different settings, with diverse groups of CYP, and younger CYP further developing it through practice and ongoing CYP involvement.

## Data Availability

The datasets used and/or analysed during the current study are available from the corresponding author on reasonable request.

## Acknowledgements

We would like to thank the CYP who generously gave their time to participate in this project and share their expert knowledge, experiences and perspectives. Their contributions were invaluable in shaping the development of this work. We would also like to thank [funder] for providing seed corn funding to support this work.

